# Implementation and economic effects of local non-pharmaceutical interventions

**DOI:** 10.1101/2022.02.10.22270783

**Authors:** Anna Godøy, Maja Weemes Grøtting

**Affiliations:** Norwegian Institute of Public Health and the Department of Health Management and Health Economics, University of Oslo; Centre for Evaluation of Public Health Measures, Norwegian Institute of Public Health

**Author notes:** Data made available by the Beredt C-19 emergency preparedness registry have been essential for the project. We are grateful to Hege M. Gjefsen, Rannveig K. Hart, Marte Strøm, Kjetil Telle, and seminar participants at the Institute for Social Research and the Norwegian Institute of Public Health for helpful discussions, suggestions, and comments. This research did not receive any specific grant from funding agencies in the public, commercial, or not-for-profit sectors. The authors have no competing interests to declare.

**Keywords:** Unemployment, mobility, COVID-19, non-pharmaceutical interventions, spillovers

## Abstract

In this paper, we analyze economic costs and consequences of local non-pharmaceutical interventions (NPIs) aimed at containing the COVID-19 pandemic. Using comprehensive data on municipal and regional policies in Norway, we implement a difference-in-differences framework identifying impacts of local NPIs from discontinuous differential shifts in outcomes following the implementation of new policies. In treated municipalities, local NPIs lead to persistent reductions in mobility, persistent increases in unemployment, and transient reductions in consumer spending. Analyses of spatial spillovers show that the implementation of local NPIs increases retail mobility in non-treated neighboring municipalities. Overall, our findings suggest that local NPIs have economic consequences for local economies and induce residents to shift their consumption of goods and services to neighboring municipalities.

## 1 Introduction

Non-pharmaceutical interventions (NPIs) have been key to managing the COVID-19 pandemic. These policies may have important financial and non-monetary costs. Policymakers face a tradeoff between controlling the number of deaths and limiting the burden of containment policies (Alvarez et al. 2020). In this paper, we consider the economic costs and consequences of local NPIs. Specifically, we implement a difference-in-differences framework, estimating effects of municipal policies on mobility, consumer spending, and unemployment.

Using difference-in-differences methods to evaluate effects of COVID-19 countermeasures is challenging for multiple reasons (Goodman-Bacon & Marcus 2020). First, the timing of policies is likely a response to COVID-19 incidence. This policy endogeneity could impact analyses of economic outcomes if higher confirmed incidence has an independent effect on economic behaviors. Second, localities may experience multiple treatments: localities may implement a social distancing mandate when rates are rising, relax the rules when incidence is lower, then re-instate the mandate later if contagion levels go back up. Third, the localities that implement NPIs do so at different times; as a result, regression difference-in-differences may yield biased estimates when treatment effects vary over time (Baker et al. 2021, Callaway & Sant’Anna 2020, Goodman-Bacon 2021). Fourth, local NPIs may have spillovers to neighboring municipalities, leading violations of the stable unit treatment value assumption (SUTVA) that underlies difference-in-differences analysis.

In this paper, we implement a stacked regression estimator (Sun & Abraham 2020, Baker et al. 2021) as implemented in Cengiz et al. (2019). This model compares outcomes in treated municipalities with outcomes in a set of clean controls, defined as municipalities that do not implement new NPIs during the entire event window. By exclusively leveraging sub-national policy variation, we ensure that the treated municipalities and the municipalities in the comparison group face the same national mandates at any given time. Our approach allows for multiple events per locality, and for treatment effects to vary over time. Moreover, these models let us assess spatial spillovers by assigning treatment status to neighboring municipalities.

Our preferred empirical specification is a set of event study regressions, estimating differential changes in outcomes over time. New NPIs are typically implemented in response to a gradual increase in observed COVID-19 incidence. To the extent that these are effective in reducing contagion, confirmed incidence would likely be gradual and occur with a lag. Conversely, mandated changes in behaviors, and economic impacts of such changes, are likely to happen more abruptly. Discontinuous shifts in the estimated event time coefficients thus point to an independent effect of NPIs on economic outcomes.

We have three main findings. First, we show that local NPIs are typically implemented as a response to high and rising rates of confirmed COVID-19 incidence. Second, we find that local NPIs significantly affect mobility patterns and economic outcomes in the treated municipalities. Our models find abrupt, persistent reductions in mobility, persistent increases in unemployment, and transient reductions in consumer spending. Third, our analysis of spatial spillovers shows that the impact of local NPIs is not limited to the treated municipalities. Rather, we find that retail mobility increases in neighboring municipalities that do not themselves introduce any NPIs during the event window. Overall, our findings suggest that local NPIs induce residents to shift their consumption of goods and services to neighboring municipalities.

Our findings contribute to the rapidly growing literature on the social and economic consequences of the pandemic and its countermeasures. Overall, these studies find mixed results on the effects of local NPIs: while social distancing mandates have been found to significantly shift mobility, studies also find that the effects of the mandates themselves may be limited relative to the voluntary behavioral changes brought about by the pandemic itself (Alexander & Karger 2021, Cronin & Evans 2020, Courtemanche et al. 2020, Goolsbee & Syverson 2021, Allcott et al. 2020, Sears et al. 2020). Most of these studies examine effects of NPIs in relatively high-incidence contexts, such as the US or other European countries with more COVID-19 related deaths. In this paper, we provide evidence of the role of NPIs in a lower incidence context, where COVID-19 fatality rates have remained relatively low throughout the pandemic. On the one hand, people may be less likely to voluntarily limit their economic and social activities when perceived risks of death or serious illness are lower; in which case the relative impact of mandates on consumer behavior and the economy may be greater. On the other hand, the content of the NPIs is typically less stringent and the lower incidence could lead to lower compliance with regulations^1^.

Our paper also contributes to the literature on spatial spillovers of local NPIs (Holtz et al. 2020, Elenev et al. 2021). The paper most similar in spirit to our own, (Elenev et al. 2021) find that stay-at-home mandates reduce mobility in neighboring counties. This result contrasts with our finding that local NPIs increase mobility in neighboring municipalities, consistent with residents substituting consumption to untreated localities. This difference could reflect the different contexts of the two studies, moreover, the NPIs we study in the present paper are implemented against the backdrop of a national policy response.

The rest of the paper is organized as follows: section 2 gives an overview of our empirical strategy. In section 3, we present an analysis linking the adoption of local NPIs to trends in confirmed incidence. Section 4 presents our analysis of the economic impacts of local NPIs, and section 5 concludes.

## 2 Empirical strategy

The impact of the COVID-19 pandemic in Norway has been geographically uneven. Local and regional policy variation has allowed policymakers to manage contagion while limiting the economic burdens of COVID-19 countermeasures. We leverage this variation to implement a difference-in-differences framework, where we estimate effects of the NPIs by comparing changes in outcomes in treated and non-treated municipalities over time.

Below, we present our empirical strategy in detail. First, we briefly review relevant institutions and background. Second, we outline the sample construction and present summary statistics of the sample. Third, we present our formal econometric models.

### 2.1 Institutions and background

The first confirmed case of COVID-19 in Norway was reported on February 27. On March 12, 2020, the Norwegian government implemented the most radical public measures in peace time. This immediate national policy response included the closing of schools and non-essential businesses and the prohibition of cultural and sporting events and all indoor recreational activities. Starting in May 2020, the lockdown was gradually lifted and low infection rates throughout September 2020 kept mandates at a minimum until October 2020, when infection rates surged.

Our paper covers the period from November 2020 to September 2021. During this period, there were no national restrictions on in-person instruction in universities, schools, preschools or daycare facilities, no national in-person retail restrictions, and no national mask mandate. For shorter periods, national prohibition of larger private gatherings and the serving of alcohol were in effect.

COVID-19 tests were widely available throughout our study period. Vaccines, on the other hand remained limited. The first vaccine was administered on December 27, 2020. Initially, vaccines were restricted to nursing home residents and selected groups of healthcare workers. By the end of April 2021, 23% had received at least one shot, while 6% were fully vaccinated. By October, 2021, the end of our sample period, 74% of the population had received at least one shot, and 64% were fully vaccinated. Due to the scarcity of vaccines, NPIs remained the most important intervention in order to contain the spread of the pandemic during most of our study period.

### 2.2 Sample construction

#### Defining events

The starting point of our analysis is data on local NPIs collected by VG, a large daily newspaper in Norway. Beginning November 19th 2020, this data represents the most comprehensive source of local and national NPIs.^2^ We focus on policies that were implemented between December 17, 2020 and September 3, 2021^3^.

In our analysis, we analyze outcomes in a 6 week window around the implementation of local NPIs. We restrict our sample to new NPIs - specifically, we restrict our sample to events where no new mandates, prohibitions, or closures were implemented in the 21 days immediately preceding the event. This restriction allows for a clean analysis of pre-trends. At the same time, we may exclude some municipalities where NPIs are updated more frequently. We make no similar restrictions on policy changes after the initial event, as these are possibly endogenous. Similarly, we do not require a minimum duration of the policy, rather, we keep the event window fixed at plus minus three weeks for all events.

The final sample includes 511 events. For each event *s*, we construct a cohort-specific estimation sample consisting of treated municipalities (new local intervention at time *s*) and clean controls (no new local interventions in an 6 week window around *s, s −* 21, …, *s* + 20). We then stack these date cohorts to obtain a balanced data set with 21 days prior to and 21 days post the intervention date. These stacked samples comprise our estimation sample.

#### Outcome variables

We construct a municipality by day (week) dataset with our outcome variables. Our analysis covers three broad categories of outcomes: (1) COVID-19 health and test outcomes, (2) behaviors, as proxied by mobility patterns, and (3) financial and economic outcomes. Data on COVID-19 related outcomes are obtained from the Emergency preparedness register for COVID-19 (Beredt C19). This is a comprehensive database of registers established to give the Norwegian Institute of Public Health an ongoing overview and knowledge of prevalence and consequences of the COVID-19 pandemic in Norway. It comprises individual-level data from a set of linkable administrative registers, including daily updated records on PCR tests and test results (MSIS) and from the Norwegian Patient Register (COVID-19 related hospitalizations), as well as the National Population Register and sociodemographic registers, including information on income, education, country of birth and housing conditions. Individuals are linked across the registers and to municipalities using unique (de-identified) personal identifiers. These data are used to construct a municipality level dataset with daily numbers of tests, confirmed cases, and hospitalizations.^4^

To assess effects of local NPIs on mobility behavior, we use two sources of data. First, we access mobility data from Google COVID-19 mobility reports (Google 2021).^5^ These reports contain daily municipality level data on mobility trends by category. We include data on mobility related to workplaces and to retail and recreation (restaurants, malls, museums, cinemas etc. excluding groceries and pharmacies).^6^ Data is missing for some of the smaller municipalities. As a consequence, mobility is observed only for a subset of events. Daily mobility is measured relative to a baseline value for the same day of the week, where the baseline is the median value for the corresponding day of the week during the period Jan 3-Feb 6, 2020.

The second mobility data source is all credit/debit card transactions of services and inperson retail for the 1.3 million private customers of DnB, a large Norwegian bank, which constitutes a market share of 26%. These data has the advantage of capturing more municipalities. However, the data is available only on a weekly basis. In addition, the data is grouped by cardholders’ municipality of residence, thus, we are not able to identify where transactions take place. Unlike the Google mobility data, which is a measure of how much mobility there is to a certain location in the municipality, the card data is therefore a measure of how much the population in the municipality moves. This distinction is useful for our analysis of spillovers.

Finally, to capture the economic consequences of local NPIs we include the following two outcomes: registered unemployment and consumer spending. For unemployment, we use publicly available data on the number of unemployed job-seekers measured each Tuesday, published by the Labor and Welfare Administration (NAV). The data allow us to distinguish between full time and part time unemployment, as well as temporary and permanent unemployment. To measure consumer spending, we use data on all credit/debit card payments from DnB described above.

#### Descriptives

Table 1 presents summary statistics of the estimation sample. The treated municipalities have higher rates of COVID-19 testing, incidence, and hospitalization relative to the comparison group.^7^ Treated municipalities tend to be larger and have a higher share of residents born abroad, higher educational attainment, and higher share of crowded housing.

**Table 1:** Summary statistics

|  | (1)<br>All | (2)<br>1+ mandate | (3)<br>No mandate |
| --- | --- | --- | --- |
| Incidence (per 100,000) | 4.080<br>(8.955) | 8.284<br>(14.22) | 4.020<br>(8.843) |
| Hospitalizations (per 100,000) | 0.131<br>(0.892) | 0.279<br>(1.355) | 0.128<br>(0.883) |
| Tests (per 100) | 254.2<br>(208.6) | 348.9<br>(237.2) | 252.8<br>(207.8) |
| Age | 41.81<br>(2.514) | 40.61<br>(2.282) | 41.82<br>(2.513) |
| Foreign born | 0.135<br>(0.0539) | 0.175<br>(0.0653) | 0.134<br>(0.0535) |
| Higher ed | 0.312<br>(0.0895) | 0.381<br>(0.110) | 0.311<br>(0.0888) |
| Cramped housing | 0.0897<br>(0.0356) | 0.113<br>(0.0456) | 0.0893<br>(0.0353) |
| Low income | 0.104<br>(0.0210) | 0.114<br>(0.0268) | 0.104<br>(0.0209) |
| Population (in 1000s) | 74.47<br>(180.9) | 184.8<br>(266.5) | 72.89<br>(178.8) |
| Observations | 1740256 | 9520 | 1730736 |

Population-weighted averages; sd in parenthesis.

Appendix table A1 summarizes the local NPIs in the estimation sample, together with a comparison sample of all local NPIs that were implemented during the sample period. The distribution of NPIs across types of polices and across policy areas is fairly similar in both samples, indicating that the sample NPIs are representative of the NPIs that were in place during the analysis period. There is considerable variation in the timing of events (see appendix figure A1). While we find one substantial hike in early January 2021 and a smaller hike towards the end of March, 2021, local NPIs were implemented throughout the sample period.

Appendix figure A2 shows the spatial distributions of the sample events, together with maps showing rates of COVID-19 testing, incidence and hospitalizations. Consistent with the results in table 1, areas that have been more impacted by COVID-19 have greater number of NPIs. At the same time, the figure shows that NPIs have been implemented in all regions of the country; they are not restricted to the capital region or larger cities.

### 2.3 Econometric models

We estimate the following event study regression model:

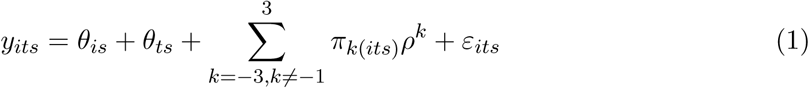

Here, *y*_*its*_ is the outcome in municipality *i*, week *t*, and event *s. π*_*k*_ is a set of event time variables, indicating that *k* weeks have passed since the implementation of the mandate. *θ*_*is*_ and *θ*_*ts*_ are event-specific fixed effects for municipality and calendar time. The calendar time fixed effects will absorb national trends in incidence, as well as any changes in NPIs at the national level.

The primary parameters of interest are the *ρ*^*k*^ attached to the event time dummies. These coefficients will capture the differential changes in outcomes in treated municipalities relative to the comparison group over the event window and relative to the week before, defined as the 7 day-period preceding the mandate. The coefficients attached to event time *k < −*1 indicate estimated pretrends, while the coefficients for *k >* 0 describe how outcomes change after the policy is implemented. Discontinuous shifts in the estimated *ρ*^*k*^ at time 0 or shortly after point to a causal effect of the mandates themselves.

To summarize the model, we also estimate the following specification, grouping event time coefficients into the following week groups: [*−*3, *−*1], 0, and [1, 2], from hereon referred to as *PRE, DURING*, and *POST*, respectively. This specification is given by:

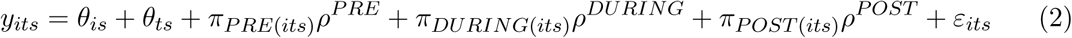

## 3 COVID-19 incidence and the implementation of local NPIs

Panel (a) in figure 1 plots average rates of testing, incidence, and hospitalizations in the 6 week window around the implementation of new local NPIs. Policies are implemented on day 1 in week 0. Consistent with the summary statistics in appendix table 1, treated municipalities have higher rates of testing, incidence, and hospitalization than untreated municipalities, but the differences are smaller between the treated and untreated neighbours compared to the full comparison group. In the weeks leading up to implementation, the gap in incidence between treated and comparison groups grows, relative to at baseline (t = –3) indicating that local NPIs are implemented as a response to increasing contagion. After the policy is implemented, the gap shrinks, with a particularly sharp decrease in the first week after the NPI took effect. As the same holds for testing, we cannot exclude that reductions in incidence are caused by reduced rates of testing, however, hospitalizations are also projecting a steep decline, indicating that the underlying prevalence is also falling. At three weeks after implementation, hospitalization rates in the treated municipalities have fallen relative to all “clean controls” and is almost similar to that of its “clean control” neighbours.

**Figure 1:**
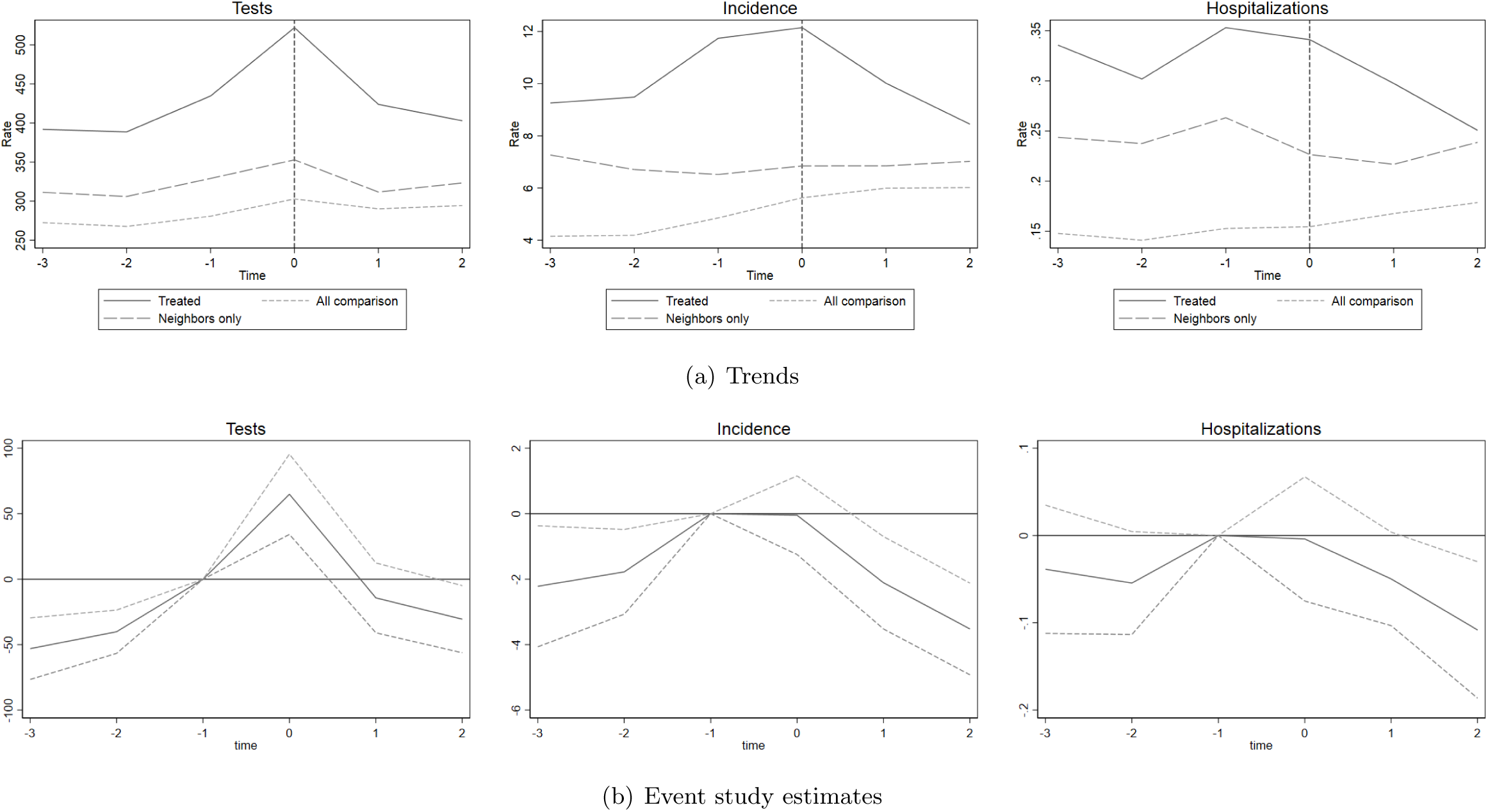
COVID-19 and the implementation of local NPIs *Note:* Panel (a) plots average rates of testing, incidence and hospitalizations in the 6 week window around the implementation of new local NPIs. “All comparison” includes all clean control municipalities (no new mandates, prohibitions or closures in the event window). “Neighbors only” includes only municipalities that are clean controls contiguous to the treatment municipalities. The averages are population-weighted. Panel (b) shows the event study estimates of equation (1) with 95% confidence intervals. The estimates are population-weighted, and standard errors are clustered at the municipality level.

Panel (b) in figure 1 presents our estimated event study models of the effects of local NPIs on testing, incidence and hospitalizations. Our models find evidence of pre-trends in test rates and incidence, which is to be expected as the purpose of local NPIs is to reduce incidence. Both increase near linearly, reaching their peak during the week the policy is introduced (NPIs are introduced on day 1 of this week). We find no significant pre-trends in hospitalizations, suggesting that local NPIs are not preceded by a surge in serious cases. After implementation, i.e. weeks 2 - 3 (marked as 1 and 2 in the graphs), there are statistically significant reductions in confirmed incidence and hospitalization rates. The reductions amounts to about 27% and 24%, respectively, relative to pre-intervention levels (see appendix table A2).

To sum up, these findings suggests that local NPIs are implemented as a response to increasing confirmed incidence and not increasing hospitalizations. Due to the endogeniety of the introduction of local NPIs with respect to incidence (also evident by the significant pre-trends), our models can not answer to the causal effects of local NPIs on contagion. However, the results on hospitalizations indicate that the local NPIs reduced the prevalence of serious cases.

## 4 Effects on mobility and the economy

### 4.1 Mobility and the economy

Our event study estimates of mobility and economic outcomes in the treated municipalities are presented in figure 2; table 2 presents the corresponding estimates from equation (2). The first four panels show effects on mobility. Panel (a) plots estimated models of workplace and retail mobility obtained from Google mobility dashboards, while panel (b) plots estimated models for in-person retail transactions and service transactions based on credit card data. The corresponding trend plots are available in appendix figure A3.

**Figure 2:**
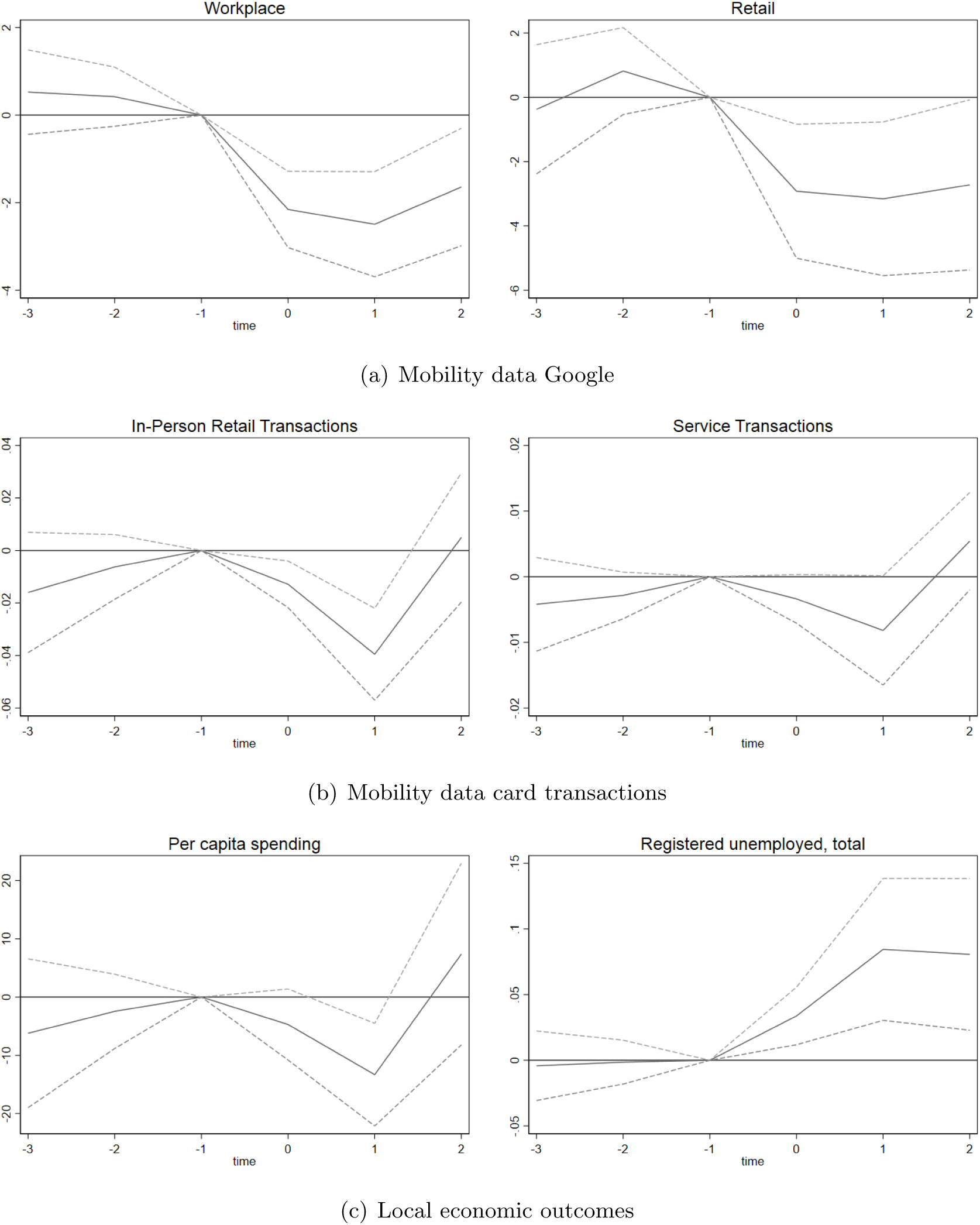
NPIs, mobility and local economies *Note:* This figure shows the event study estimates of equation (1) with 95% confidence intervals. The estimates are population-weighted, and the standard errors are clustered at the municipality level.

**Table 2:** Mobility and the economy in treated municipalities

|  | (1)<br>Workplace | (2)<br>Retail | (3)<br>In-person<br>transactions | (4)<br>Service<br>Transactions | (5)<br>Per capita<br>spending | (6)<br>Unemployment |
| --- | --- | --- | --- | --- | --- | --- |
| PRE | 0.474<br>(0.323) | 0.222<br>(0.746) | -0.0111<br>(0.00810) | -0.00352<br>(0.00236) | -4.34<br>(4.39) | -0.00276<br>(0.0103) |
| DURING | -2.15*** | -2.92*** | -0.0129*** | -0.00337* | -4.72 | 0.0338*** |
|  | (0.431) | (1.01) | (0.00451) | (0.00188) | (3.10) | (0.0111) |
| POST | -2.07*** | -2.94** | -0.0173* | -0.00137 | -2.97 | 0.0826*** |
|  | (0.587) | (1.16) | (0.00904) | (0.00349) | (5.55) | (0.0282) |
| N | 65,142 | 15,666 | 743,544 | 388,764 | 404,676 | 731,640 |
| Pre-Mean | -29.9 | -21.4 | 1.32 | 0.235 | 656 | 4.66 |
| Relative change | - | - | -0.0131 | -0.00582 | -0.00453 | 0.0177 |

*Note:* The table presents estimates of equation (2). All models include municipality-by-event and day-by-event fixed effects. Estimates are population-weighted; standard errors are clustered at the municipality level.

We find no significant pre-trends in any of the mobility outcomes, suggesting that the mobility outcomes would have trended in parallel in absence of the introduction of new local NPIs. When the local NPIs are introduced, there us a discontinuous downward shift in mobility for all mobility outcomes. The event study graphs in panel (a) show statistically significant and persistent reductions in mobility to workplace and retail locations.

These reductions in mobility arrive in the context of already low mobility levels in the treated municipalities. On average, workplace and retail mobility rates in the pre-implementation period are down by 30% relative to pre-pandemic baselines. The implementation of NPIs then reduce mobility in these two categories by an additional 2% and 3% respectively^8^.

Panel (b) presents the corresponding analyses of in-person card transactions.^9^ For both the number of in-person transactions and the number of service transactions, we find significant shifts after the policy is implemented. However, while the shifts in Google mobility data were persistent, the reduction in numbers of card transactions appears to be temporary: by the end of the event window, the estimated event time coefficients are not statistically significantly different from zero. As a consequence, the overall impact of NPIs on transactions in the 3 weeks after implementation is limited: relative to pre-implementation means, in-person retail and service transactions drop by an average of 1.3% and 0.6%, respectively (see table 2).

In the results in figure 2, event time is defined in weeks relative to implementation. However, as the Google mobility data is available on a daily basis, we also present estimates of daily mobility responses using +/- 10 days as estimation windows. These results are presented in appendix figure A4. As in the weekly models, the daily models show a discontinuous shift in mobility following the implementation of new local NPIs. Moreover, these results show that the effects on mobility were immediate.

To summarize, the introduction of local NPIs is followed by a substantial reduction in mobility. While this holds for all four mobility outcomes, there is a discrepancy in that the effects using Google mobility data show persistent reductions in mobility, whereas the card transaction data show only a transient drop. As the google mobility data defines mobility based on activity in a given geographic area, while the card data defines mobility based on the card holder’s municipality of residence, what we see is a persistent drop in mobility to certain places, but people’s spending patterns are only temporarily reduced. This discrepancy could reflect that people shift to less risky forms of consumption, e.g. take-away dinners instead of in-person eating; or a spatial shift where consumers seek out retail and services in neighboring (untreated) municipalities. We will return to the latter possibility in our analysis of spatial spillovers.

Returning to figure 2, panel (c) presents the effects of the local NPIs on local economies. Here, we consider two outcomes: consumer spending and registered unemployment. For consumer spending, we see no significant pre-trends before there is a significant drop in spending during the first post-intervention week. Quantitatively, the effect is modest: we estimate a reduction of about 15 NOK/day, which corresponds to about 2% relative to pre-implementation levels in the treated municipalities (column (5) table 2). In week 3, spending returns to pre-intervention levels, suggesting that the drop in consumer card purchases is temporary. Thus, the overall impact of local NPIs on consumption in the post period is close to zero (see column (5), table 2).

We have also estimated models separately for online and in-person purchases. Results from this exercise are presented in appendix figure A5 and appendix table A3. While there are no significant pre-trends in neither in-person nor online purchases, there is a slight increase in online purchases coupled with a significant drop in in-person transactions. That is, consumers appear to shift some consumption online at the implementation of local NPIs.

For unemployment, we find no evidence of significant pre-trends. In the week of implementation, registered unemployment increases slightly, before it jumps further in week 2 and stays elevated through week 3. Quantitatively, the effects are small: we estimate an increase in weeks 2 - 3 to be about 5% relative to pre-implementation levels in the treated municipalities. At the same time, these represent significant income losses for the affected workers, in particular because the increase is driven entirely by increases in fully-unemployed persons, with part-time unemployment/hours reduction basically unchanged (see appendix figure A6 and table A4).

The transient drop in consumer spending contrasts with the persistent increase in registered unemployment. One possibility is that the bounce back reflects consumers shifting away from affected businesses (e.g. bars) towards unaffected sectors (e.g. coffee shops) or to establishments in other municipalities.

### 4.2 Spillovers to neighboring municipalities

Spillovers can both through changes in cross-municipality mobility patterns of residents in treated municipalities (e.g. closing bars and restaurants lead residents in treated municipalities to seek out in-person dining in neighboring municipalities), and through changes in behaviors of residents in neighboring municipalities (e.g. mandated closures in the treated municipalities lead to residents in neighboring municipalities staying home more). To assess spillovers to neighboring municipalities, we estimate a version of the event study model where the neighboring municipalities are assigned the treatment dates of the treated municipalities, and the comparison groups consists of all “clean controls” that that are not neighboring a treatment municipality. Results from this exercise are shown in figure 3; corresponding point estimates from equation (2) are presented in appendix table A5.

**Figure 3:**
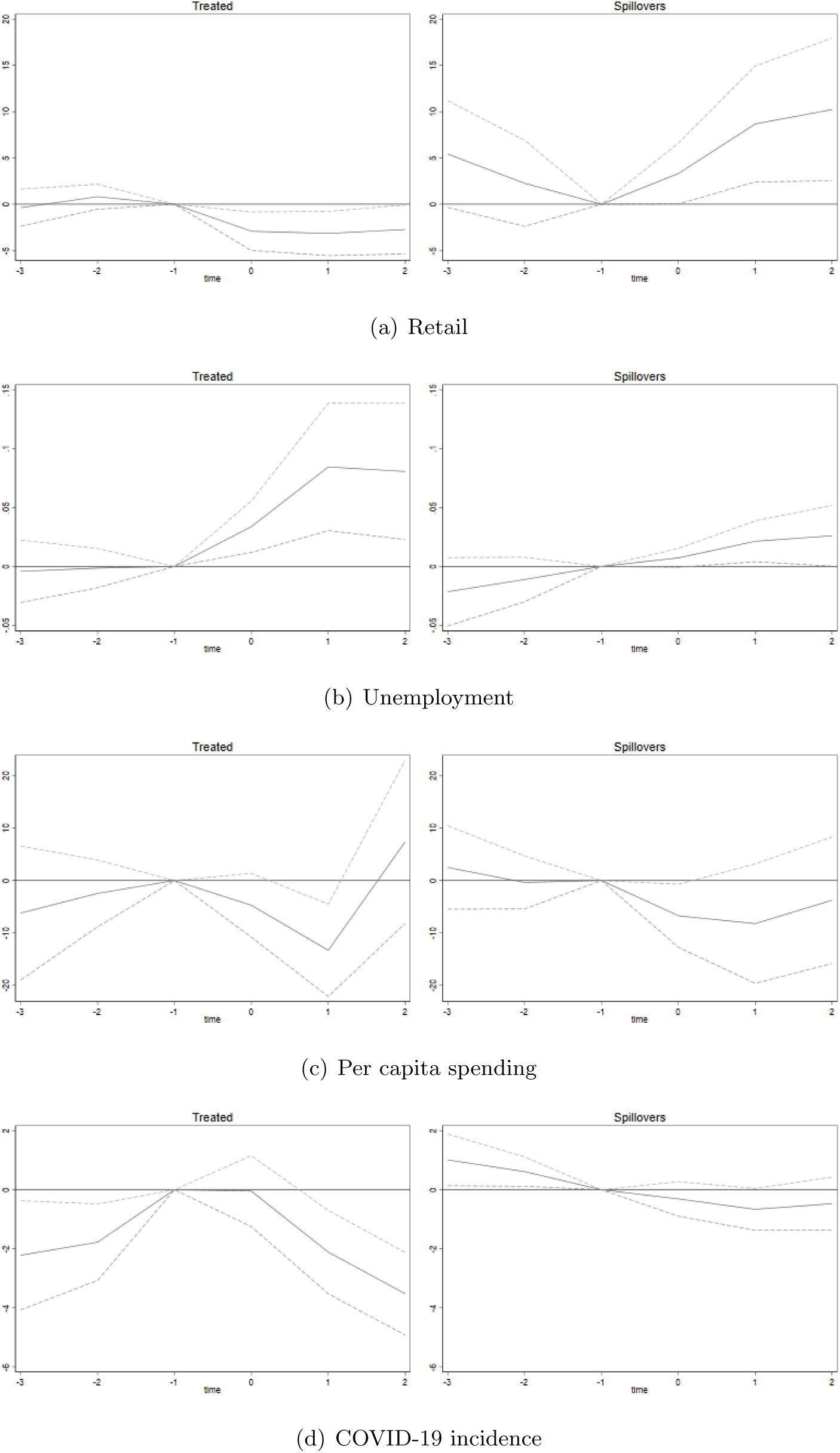
Spillovers to neighboring municipalities *Note:* This figure shows the event study estimates of equation (1) with 95% confidence intervals. The estimates are population-weighted, and the standard errors are clustered at the municipality level.

Panel (a) presents our analysis of spillovers in retail, showing estimated event study models of retail mobility in treated (our main results) municipalities to the left and neighboring municipalities to the right. When NPIs are implemented in one municipality, mobility related to retail increases in neighboring municipalities, compared to the rest of the “clean control” municipalities. Panels (b) and (c) of figure 3 show effects on unemployment and consumer spending for residents in neighbouring municipalities. Overall, we find little evidence of significant spillovers for either of these outcomes.^10^

The analysis of spillovers indicate that when local NPIs are introduced in one municipality, people travel across municipality borders. This change in mobility patterns can also explain the discrepancy between permanent increases in unemployment, but only transitory reductions in consumer spending among people living in the affected municipality found in our main results. To find out if this increase in mobility, plausibly by people from higher infection areas, also results in higher infection levels, we also analyse spillovers in COVID-19 incidence. This does not seem to be the case, as panel (d) shows that there were no statistically significant spillovers in COVID-19 incidence.

### 4.3 Robustness

As table 1 makes clear, municipalities that implement local NPIs are systematically different from the “clean controls” comparison group. The treated municipalities tend to be significantly larger and have higher initial rates of testing, confirmed incidence, and hospitalizations. To account for these differences, we implement a propensity score reweighting approach to impose balance across treatment and comparison municipalities (Bailey & Goodman-Bacon 2015, Goodman-Bacon & Marcus 2020).

To estimate the propensity scores, we include the following pre-determined variables: log of population, the share of employed who were employed in hotels and restaurants as of March 1, 2020, municipality centrality class (six dummies), share of population with low income, share of population older than 40 years, share of population living in crowded housing, and share of population with higher education. We also include weekly rates of COVID-19 testing, incidence, and hospitalization three weeks prior to the implementation of a local NPI (Goodman-Bacon & Marcus 2020).^11^

In a related exercise, we augment our baseline specifications in equations (1) and (2) with the following covariates: municipality population and the share of employed who were employed in hotels and restaurants as of March 1, 2020. The controls are interacted with calendar time.

The re-weighted and covariate adjusted estimates are presented in figure 4. These results show that especially re-weighting, but also adding covariates, substantially increases the effects on unemployment and spending. The results from the event study are summarized in table 3 (columns (1) - (2) and (6) - (7)). The overall effect in the post period is a relative increase in unemployment of 4% (2%) for the re-weighted (covariate adjusted) model. Unlike in the main model, the overall effect on spending is negative and statistically significant. The relative change compared to pre-implementation spending is a reduction of about 2% for both re-weighted and covariate adjusted results, which is still qualitatively small. However, the effects on unemployment are not trivial.

**Figure 4:**
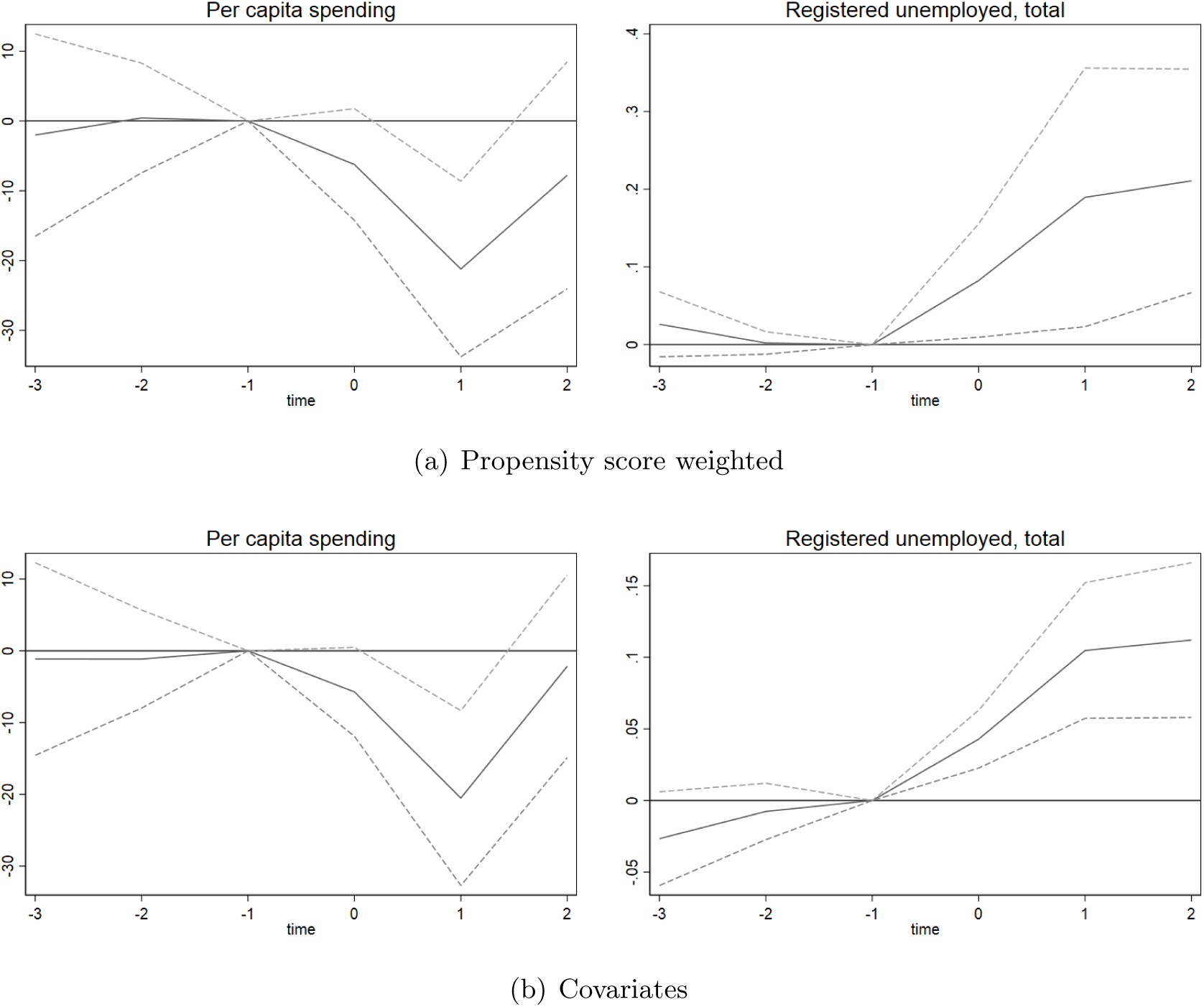
Propensity score weighted event study estimates *Note:* This figure shows the event study estimates of equation (1) with 95% confidence intervals. The estimates are propensity score and population-weighted, and the standard errors are clustered at the municipality level.

**Table 3.**
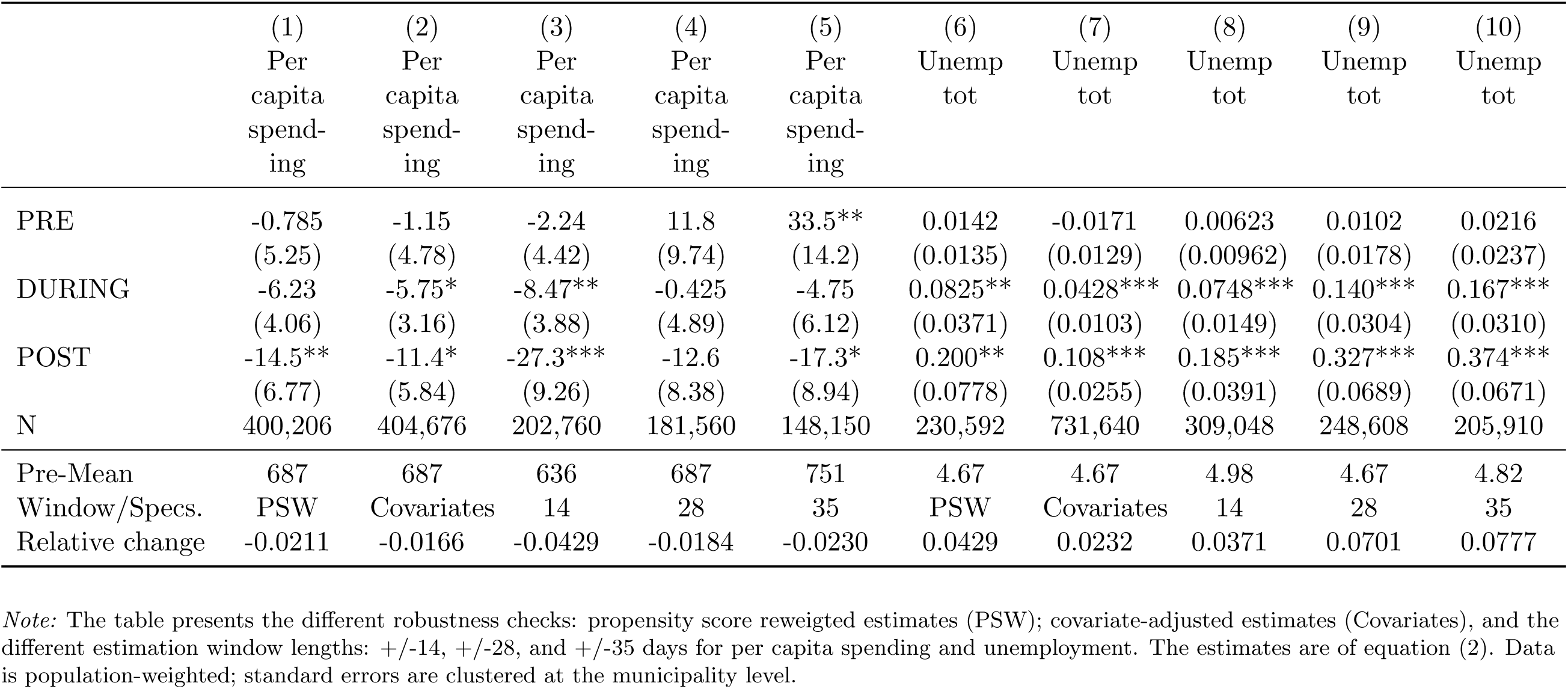
Robustness: propensity score reweighting, covariates, and event window

To assess whether our results are driven by any single events, we implement a leave-one-out approach, re-estimating the regression model of equation (2) sequentially leaving out each event from the estimation sample. Results from this exercise are shown in appendix figure A8. In this figure, results from our main specification is shown as a thick black line. We find some indication that two single events shift the magnitude of the estimated effects. Qualitatively, the pattern remains constant across estimation samples.

In our baseline models, we use a symmetric +/- 21 days window to define events. This choice is not trivial: while a longer window allows for a more careful analysis of pre-trends and treatment effects, this comes at a cost of excluding more events from the sample. A longer window imposes more stringent criteria for the comparison municipalities, potentially affecting the size and composition of the comparison group. Requiring no new local policies over a longer time will generally lead to the comparison group being made out of smaller municipalities with lower rates of COVID-19 incidence and hospitalization.

To address the robustness of our findings to the choice of window length, we re-estimate the models in equations (1) and (2) on samples where the event windows are set to +/- 14, 28 and 35 days, respectively. Appendix table A6 shows summary statistics of these samples. The shortest window, +/- 14 days with no new mandates, yields treatment and comparison municipalities that are more similar based on observables, whereas the longest window, +/- 35 days with no new mandates, causes the comparison municipalities to differ more substantially from the treated municipalities, in particular with regards to population size and rates of COVID-19 infections and hospitalizations.

The estimation results from this exercise are summarized in panels (3)-(5) and (8)-(10) in table 3.^12^ There is a larger reduction in consumption for the shortest window, but this becomes smaller when the estimation window is increased. These results are in line with event study graphs of our main results (figure 2) where there is a short term effect on spending that levels off over time. For unemployment the result is the reverse. Here, the effect increases as the estimation window increases. This is also in line with the main event study graphs for unemployment, where employment is found to continue to increase as time from implementation of the NPI passes. For event windows 28 and 35 days post implementation, unemployment increases by 0.3 and 0.4 percent points, respectively, which amounts to a relative change in unemployment of 7% and 8% compared to pre-implementation levels.

### 4.4 Discussion

In this paper, we estimate economic effects of local NPIs in Norwegian municipalities. Using data on registered unemployment and card transactions from a large Norwegian bank, we estimate a series of event study models capturing differential changes in outcomes in treated and comparison municipalities upon the implementation of new NPIs. We find that local NPIs have significant economic impacts on the treated municipalities. Immediately following implementation, there is a significant drop in consumer spending as well as a significant increase in registered unemployment. While the reduction in consumer spending is transitory, the increase in registered unemployment is persistent. We further analyse spillovers to neighboring municipalities, and show that while the implementation of a local NPI reduces mobility to retail locations in the focal municipality, it increases mobility to retail locations in neighboring municipalities that did not implement any NPIs.

Our results contrast with the findings of Elenev et al. (2021) who find that stay at home mandates reduce mobility in neighbouring counties. This difference could reflect differences in settings: their paper estimates effects of stay-at-home orders in the United States, a relatively high-incidence, high fatality setting. Meanwhile, we estimate effects of less far-reaching NPIs in a setting where incidence and fatalities have been relatively low.

The low-fatality setting could also explain the differences between our findings and the conclusions in Goolsbee & Syverson (2021) that individual choices, rather than mandates, was the primary driver of economic decline in the pandemic. In our low fatality setting, the perceived risk of death or serious illness from regular social interactions is likely lower. As a result, the fear of infection may have less relative impact on behaviors. Local NPIs, even if they stop short of a full stay-at-home mandate, may therefore have greater relative effect on mobility and economic outcomes.

To illustrate the implications of our findings, we have carried out a simple policy simulation, using the estimated 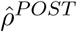 to calculate the estimated increases in registered unemployment that result from the local NPIs. Results from this exercise are presented in Appendix table A7. Our point estimates suggest that the local NPIs led to a total of 10,800 excess unemployed workers over the post-period; the large majority (more than 8,800) of these were laid off full-time.

Our data does not allow us to identify individual unemployed workers. Meanwhile, numbers from Statistics Norway show that the economic impacts of the pandemic have been highly uneven: immigrants, Norwegian-born children of immigrants, and workers with less formal education have higher rates of job loss during the pandemic (see appendix figure A10). Data from past recessions suggests that such job loss could have lasting effects on earnings, employment, health and mortality (von Wachter 2020, Oreopoulos et al. 2012). Immigrants and their descendants may be more vulnerable to these adverse outcomes due to discrimination in the labor market (Oreopoulos 2011, Midtbøen 2016, Hermansen 2013).

Taken at face value, our point estimates would imply that the NPIs averted approximately 370 confirmed cases and 10 hospitalizations. However, these numbers should be interpreted with caution given the problems with identifying causal impacts on incidence when policies are endogenous. The violation of parallel trends imply that the non-treated comparison municipalities are not a valid counterfactual for the evolution of incidence and hospitalizations absent the NPIs. In particular, the estimated pre-trends in figure 1 suggest that the true impacts on incidence and hospitalization are likely significantly larger.

## 5 Conclusions

The rapidly growing literature on the impacts of NPIs finds mixed results on the relative importance of these policies. Our analysis shows that in the low-incidence context of Norway, local NPIs have statistically significant, economically meaningful effects on local economies.

These findings have implications for further research. In particular, the literature on the lasting effects of job loss on later health and labor market outcomes suggests that the documented increases in unemployment have potentially far-reaching implications. Evaluating the long-term impacts of NPIs should remain a priority as more data becomes available.

## Data Availability

Sensitive data kept in repository at Norwegian Institute of public health

### Appendix A

**Table A1:** Types of policies in our sample and in all NPIs

|  | Sample only | All NPIs |
| --- | --- | --- |
| Prohibitions | 0.378 | 0.368 |
| Mandates | 0.568 | 0.521 |
| Closures | 0.360 | 0.430 |
| Masks | 0.327 | 0.251 |
| Leisure | 0.630 | 0.656 |
| Social contact | 0.661 | 0.620 |
| Work | 0.125 | 0.148 |
| Education | 0.229 | 0.253 |
| Travel | 0.125 | 0.146 |
| Christmas | 0.0157 | 0.0207 |
| Observations | 511 | 677 |

This table shows the distribution of local NPIs into types of policies (prohibitions, mandates, and closures) and into policy categories for our sample of NPIs (first column) and for all NPIs implemented during our study period (second column). Mean coefficients; $t$ statistics in parentheses.

**Figure A1:**
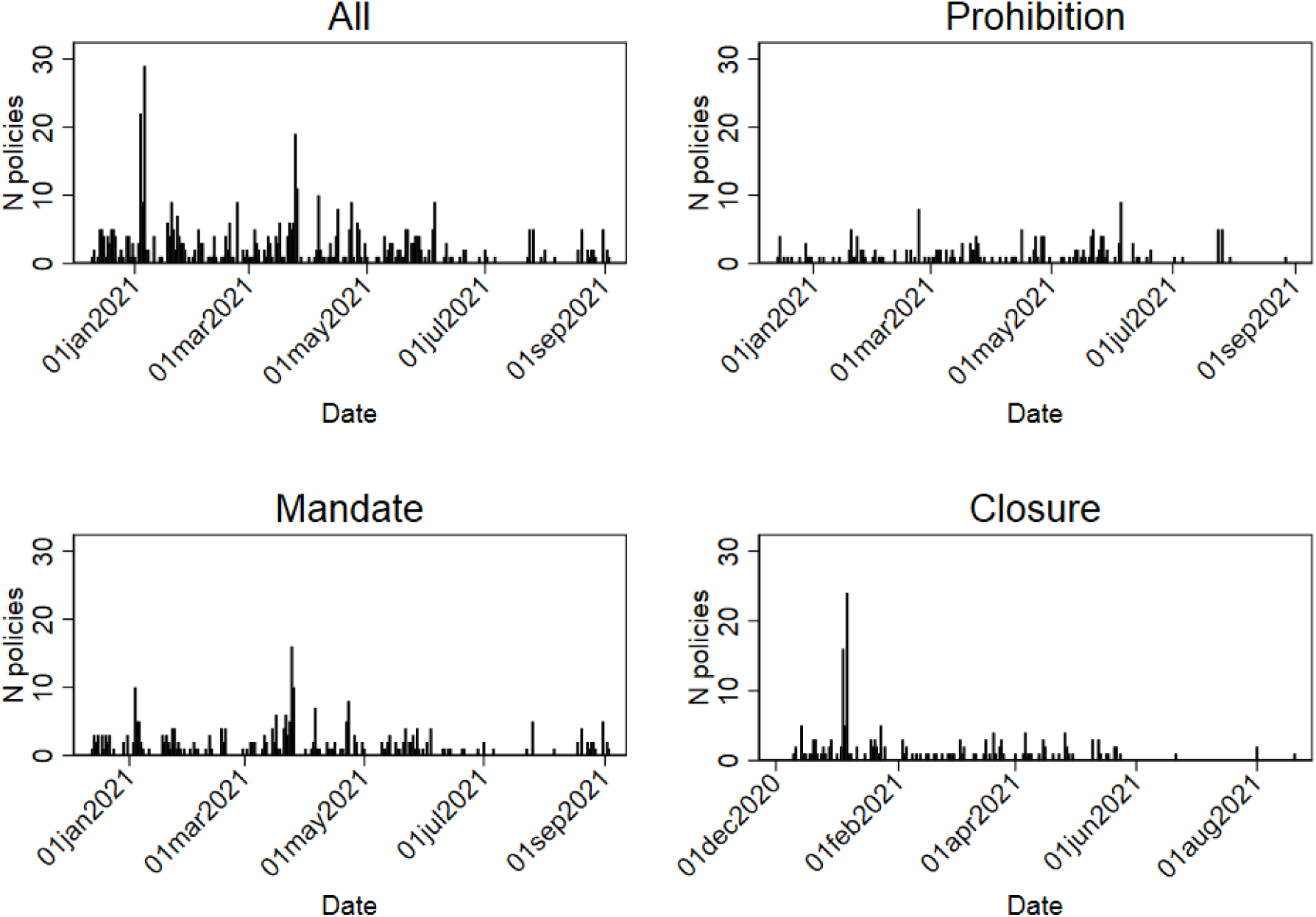
Timing of local NPIs *Note:* This figure shows daily number of new local NPIs between December 17, 2020 and Sept 29, 2021.

**Figure A2:**
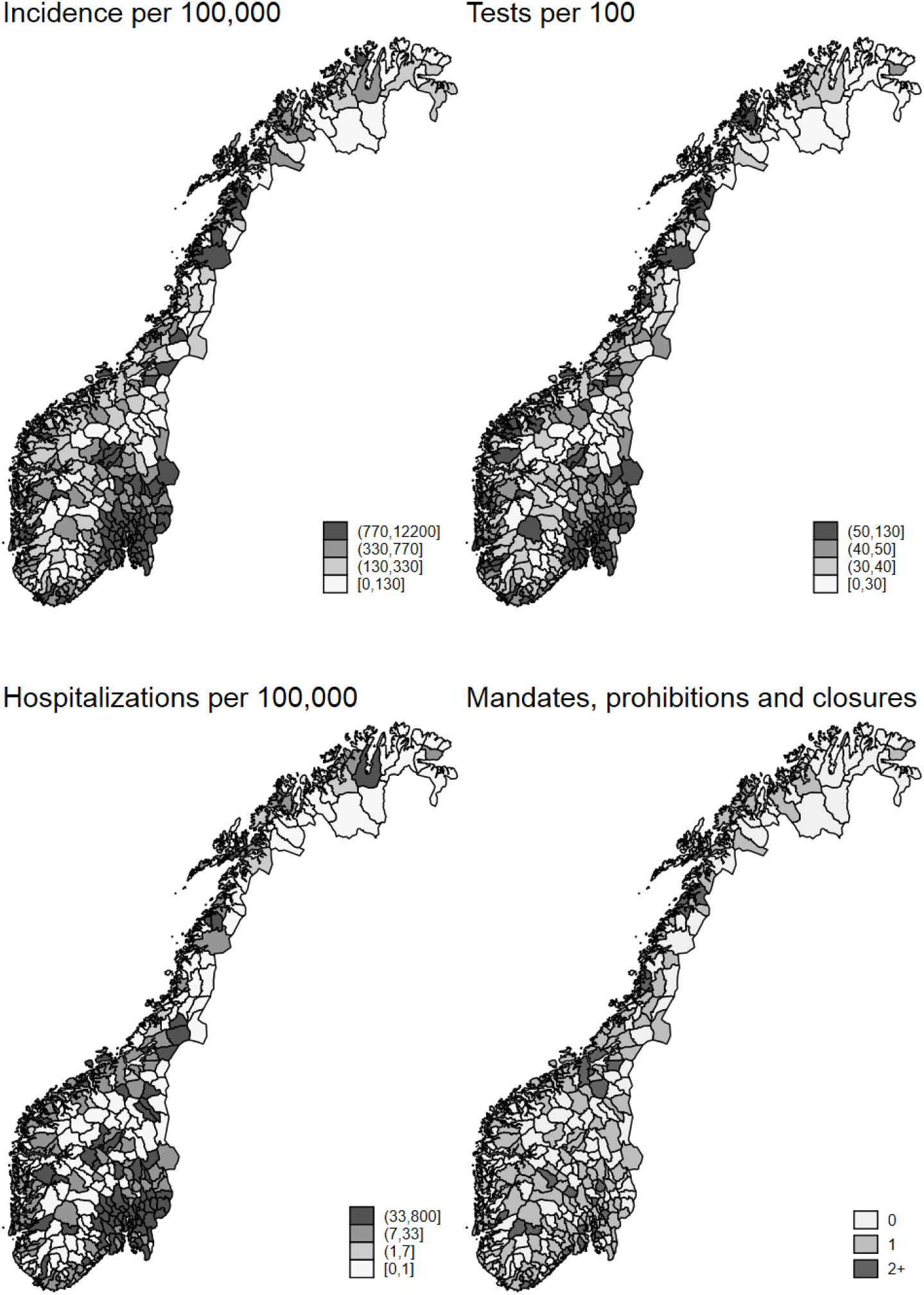
NPIs, incidence, hospitalizations, and tests *Note: This figure presents the geographic distribution of NPIs and COVID-19 tests, incidence, and hospitalizations across Norwegian municipalities*.

**Figure A3:**
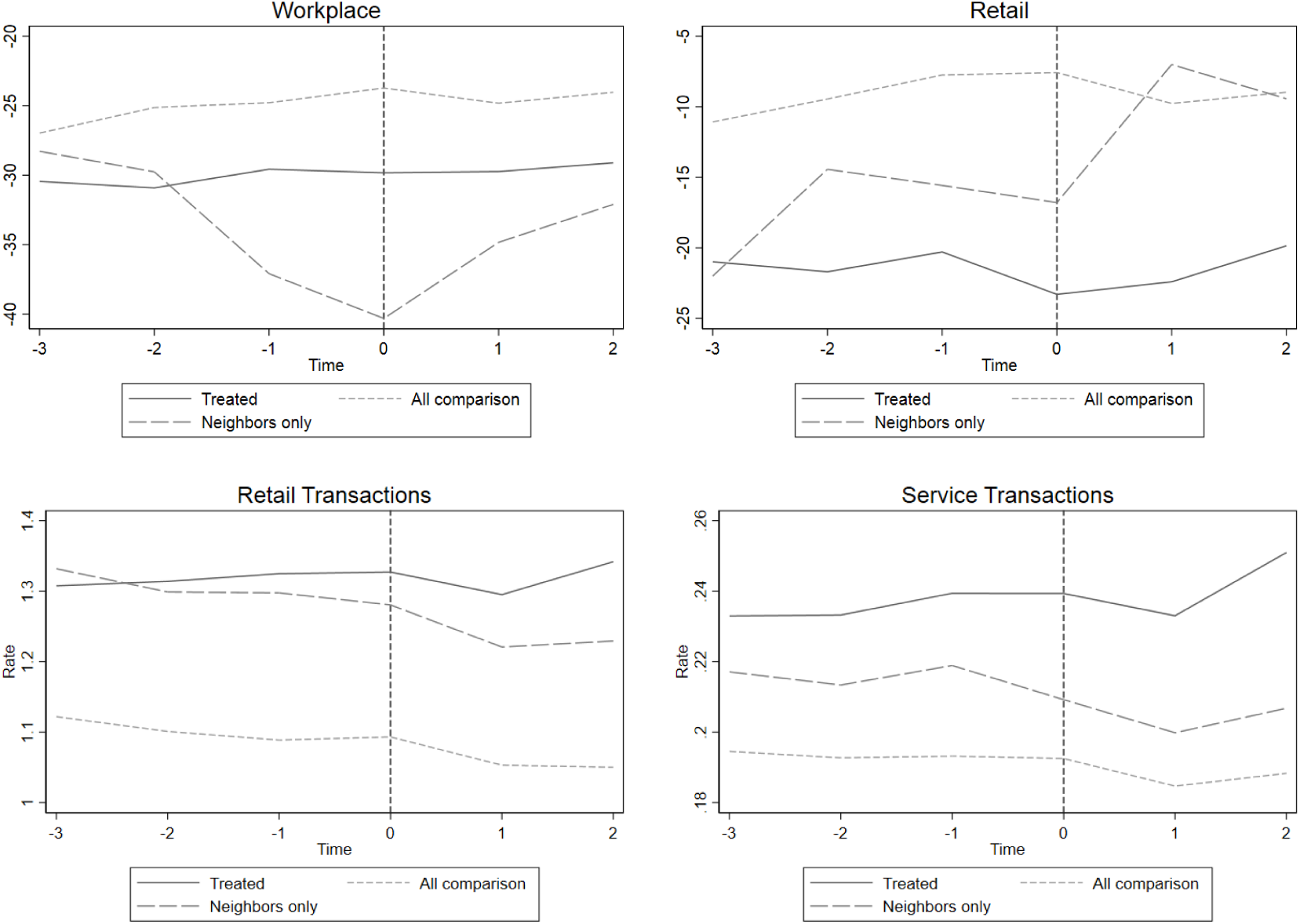
Trends - Mobility patterns *Note:* this figure show population-weighted trends in mobility. The first four graphs are from Google mobility data and the bottom two graphs are from DNB transaction data.

**Table A2:** Testing, incidence, and hospitalization Diff-in-Diff

|  | (1)<br>Incidence | (2)<br>Tests | (3)<br>Hospitalizations |
| --- | --- | --- | --- |
| PRE | -2.00**<br>(0.784) | -46.6***<br>(9.44) | -0.0465<br>(0.0307) |
| DURING | -0.0453<br>(0.611) | 64.9***<br>(15.6) | -0.00389<br>(0.0362) |
| POST | -2.81***<br>(0.655) | -22.4*<br>(12.4) | -0.0789**<br>(0.0309) |
| N | 5,121,480 | 5,121,480 | 5,121,480 |
| Pre-Mean | 10.2 | 405 | 0.330 |
| Relative change | -0.277 | -0.0553 | -0.239 |

The table presents estimates of equation (2) for the different COVID-19 outcomes. Standard errors are clustered at the municipality level.

**Figure A4:**
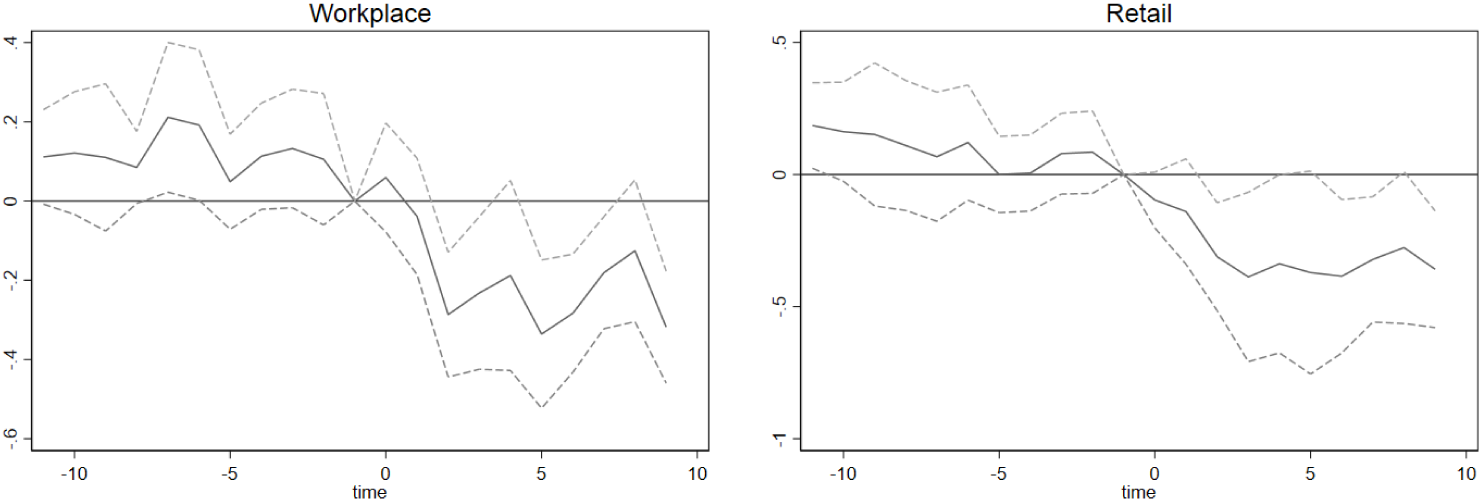
Event study estimates: days mobility *Note:* This figure shows the event study estimates of equation (1) with 95% confidence intervals and +/- 10 days estimation window. The estimates are population-weighted, and the standard errors are clustered at the municipality level. The table presents estimates of equation (2) for different types of card transactions. Standard errors are clustered at the municipality level.

**Table A3:** Spending Diff-in-Diff

|  | (1)<br>Per capita spending | (2)<br>In person | (3)<br>Online |
| --- | --- | --- | --- |
| PRE | -4.34<br>(4.39) | -2.41<br>(3.96) | -1.49<br>(1.52) |
| DURING | -4.72<br>(3.10) | -3.36<br>(3.21) | -0.108<br>(0.926) |
| POST | -2.97<br>(5.55) | -3.37<br>(6.52) | 0.375<br>(1.53) |
| N | 404,676 | 743,544 | 526,656 |
| Pre-Mean | 656 | 498 | 88.7 |
| Relative change | -0.00453 | -0.00677 | 0.00422 |

The table presents estimates of equation (2) for different types of card transactions. Standard errors are clustered at the municipality level.

**Table A4:** Registered Unemployment Diff-in-Diff

|  | (1)<br>Unemp tot | (2)<br>OU- PT | (3)<br>TL - PT | (4)<br>OU-FT | (5)<br>TL - FT |
| --- | --- | --- | --- | --- | --- |
| PRE | -0.00276<br>(0.0103) | -0.00260<br>(0.00397) | -0.00431<br>(0.00301) | 0.00133<br>(0.00530) | 0.00282<br>(0.00401) |
| DURING | 0.0338***<br>(0.0111) | 0.00521**<br>(0.00241) | 0.00385*<br>(0.00227) | 0.0150**<br>(0.00619) | 0.00969**<br>(0.00447) |
| POST | 0.0826***<br>(0.0282) | 0.00899<br>(0.00620) | 0.00705<br>(0.00493) | 0.0416***<br>(0.0131) | 0.0249***<br>(0.00839) |
| N | 731,640 | 731,640 | 731,640 | 731,640 | 731,640 |
| Pre-Mean | 4.66 | 1.28 | 0.640 | 2.11 | 0.635 |
| Relative change | 0.0177 | 0.00705 | 0.0110 | 0.0198 | 0.0392 |

The table presents estimates of equation (2). Standard errors are clustered at the municipality level. OU-PT: ordinary unemployed, part time; TL-PT: temporarily layoff, part time; OU-FT: ordinary unemployed, full time; TL-FT: temporarily layoff, full time.

**Table A5:** Spillovers to neighboring municipalities

|  | (1)<br>Retail | (2)<br>Unemployment | (3)<br>Per capita<br>spending | (4)<br>Incidence |
| --- | --- | --- | --- | --- |
| PRE | 3.85<br>(2.44) | -0.0163<br>(0.0115) | 1.06<br>(3.02) | 0.814**<br>(0.319) |
| DURING | 3.32**<br>(1.56) | 0.00729*<br>(0.00407) | -6.72**<br>(3.06) | -0.310<br>(0.298) |
| POST | 9.47***<br>(3.17) | 0.0237**<br>(0.0100) | -5.99<br>(5.83) | -0.566<br>(0.389) |
| N | 3,570 | 728,574 | 608,952 | 5,100,018 |
| Pre-Mean | -29.6 | 4.26 | 656 | 6.83 |
| Relative change | -0.319 | 0.00556 | -0.00913 | -0.0829 |

The table presents estimates of spillover effects in mobility to retail locations, unemployment rate, per capita spending of the municipality inhabitants, and COVID-19 incidence. Estimates are of equation (2), and standard errors are clustered at the municipality level.

**Figure A5:**
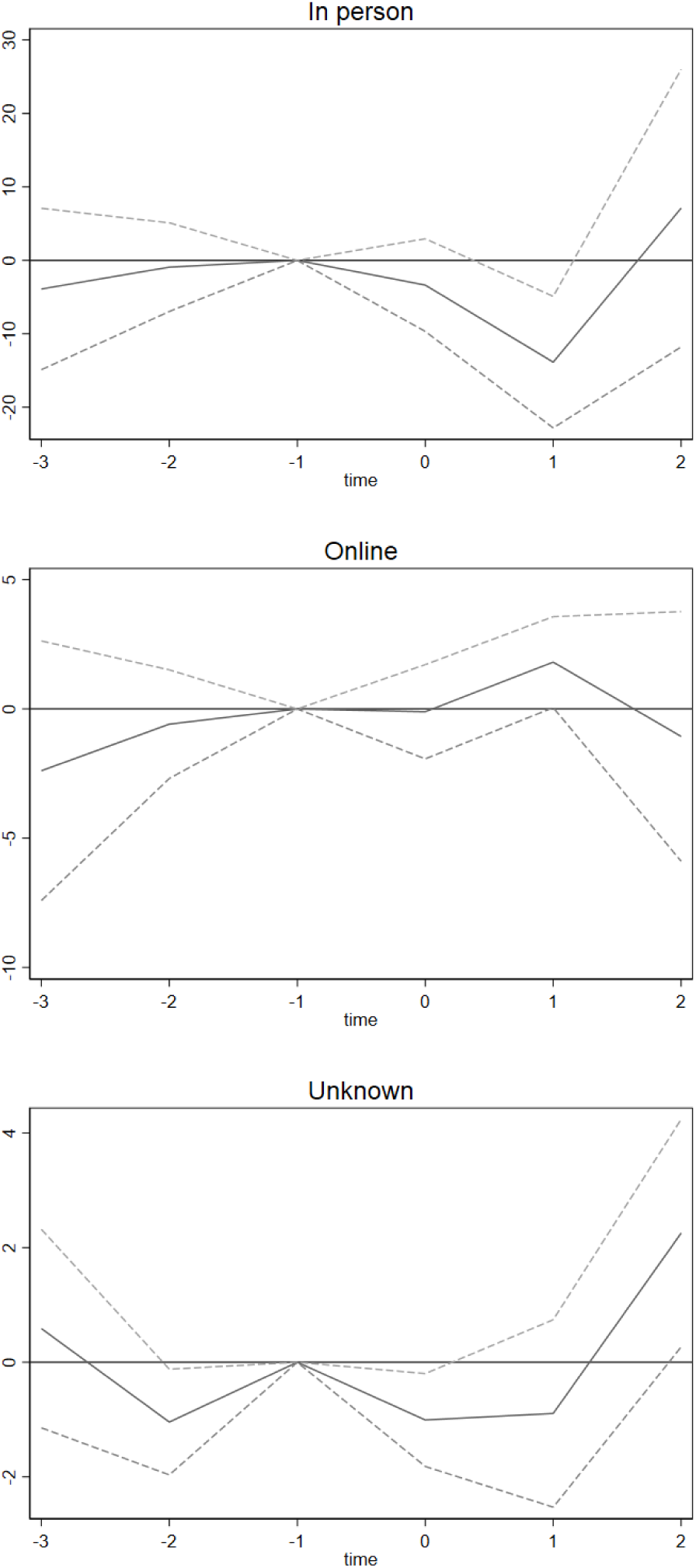
Consumer spending - online vs in person *Note:* This figure shows the event study estimates of equation (1) with 95% confidence intervals using the card data and different types of transactions. The estimates are population-weighted, and the standard errors are clustered at the municipality level.

**Figure A6:**
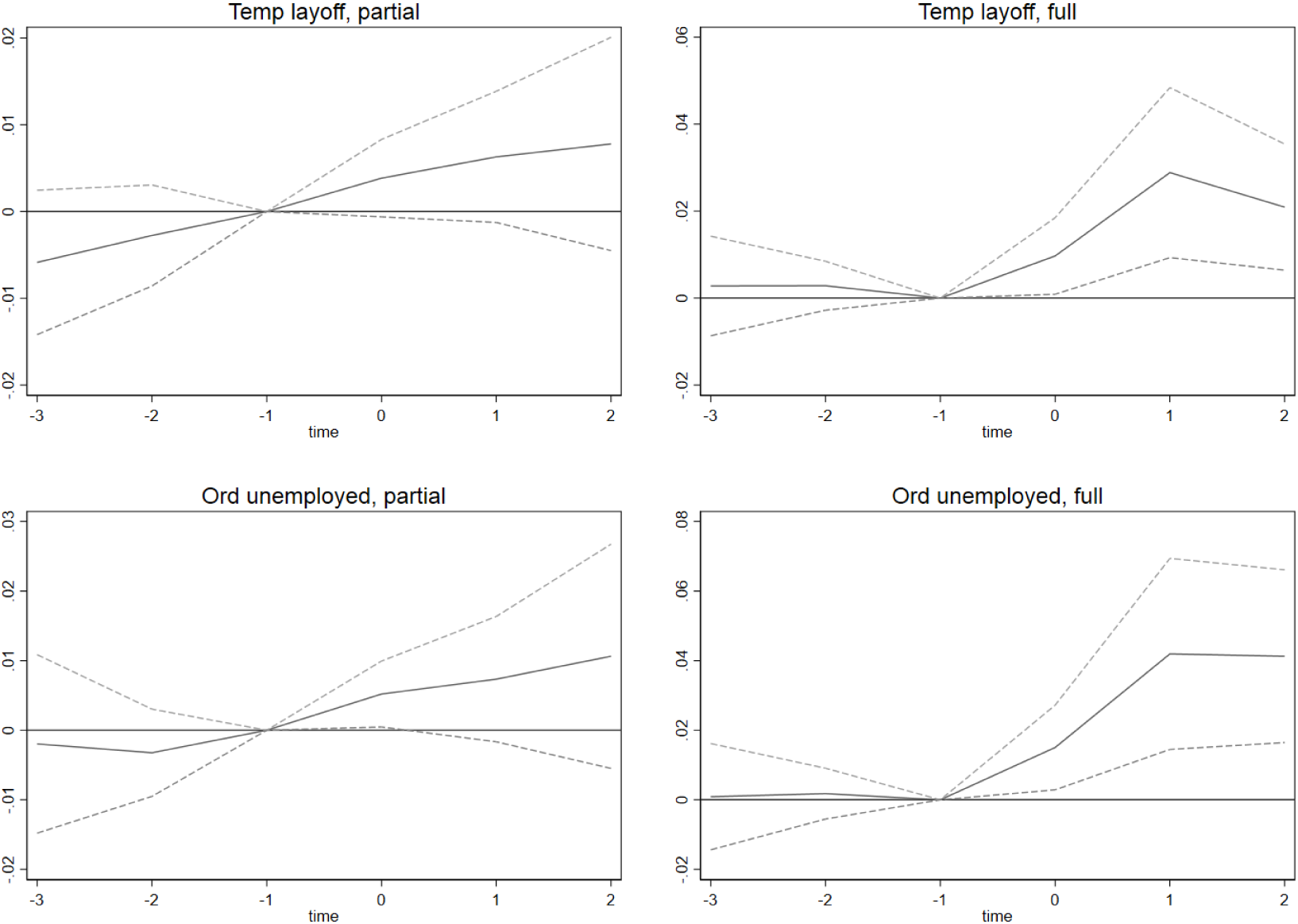
Unemployment by category *Note:* This figure shows the event study estimates of equation (1) with 95% confidence intervals using the unemployment data for different types of unemployment categories. The estimates are population-weighted, and the standard errors are clustered at the municipality level.

**Figure A7:**
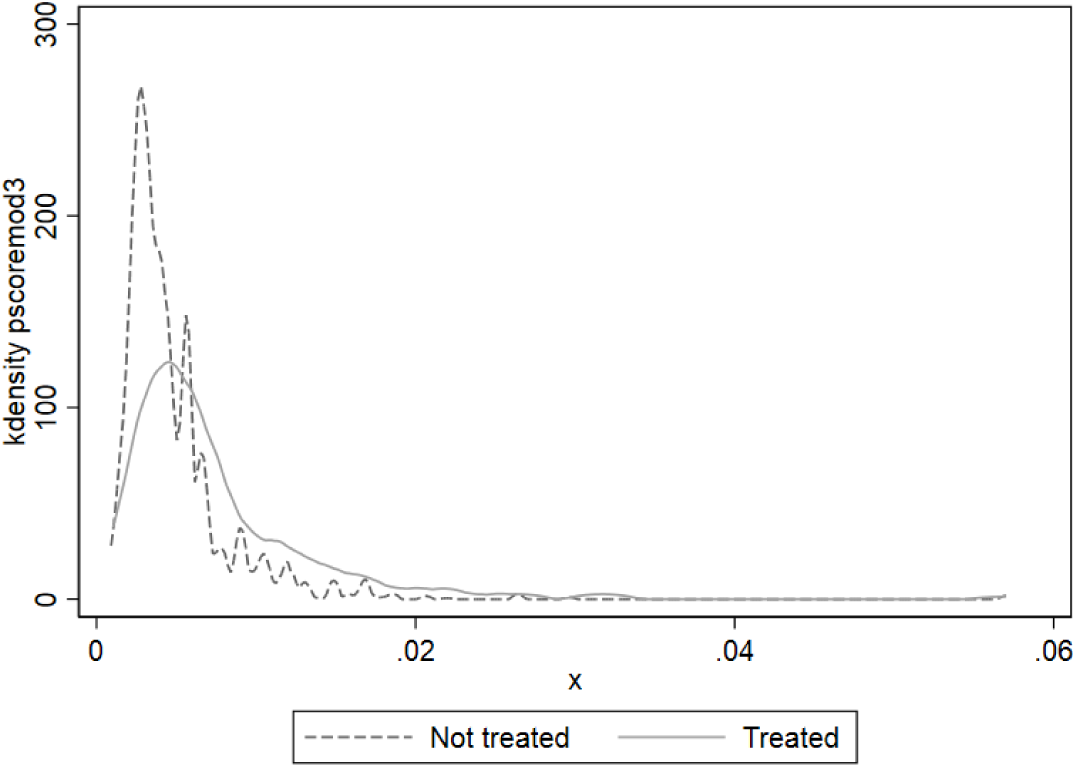
Common support propensity weights *Note:* The graph shows the common support graphs made by our stacked sample, where the controls can appear multiple times as they can be controls for multiple events. The propensity scores are calculated using the following pre-determined variables: log of population, the share of employed who were employed in hotels and restaurants as of March 1, 2020, municipality centrality class (six dummies), share of population with low income, share of population older than 40 years, share of population living in crowded housing, and share of population with higher education, as well as weekly rates of COVID-19 testing, incidence, and hospitalization three weeks prior to implementation of a local NPI

**Figure A8:**
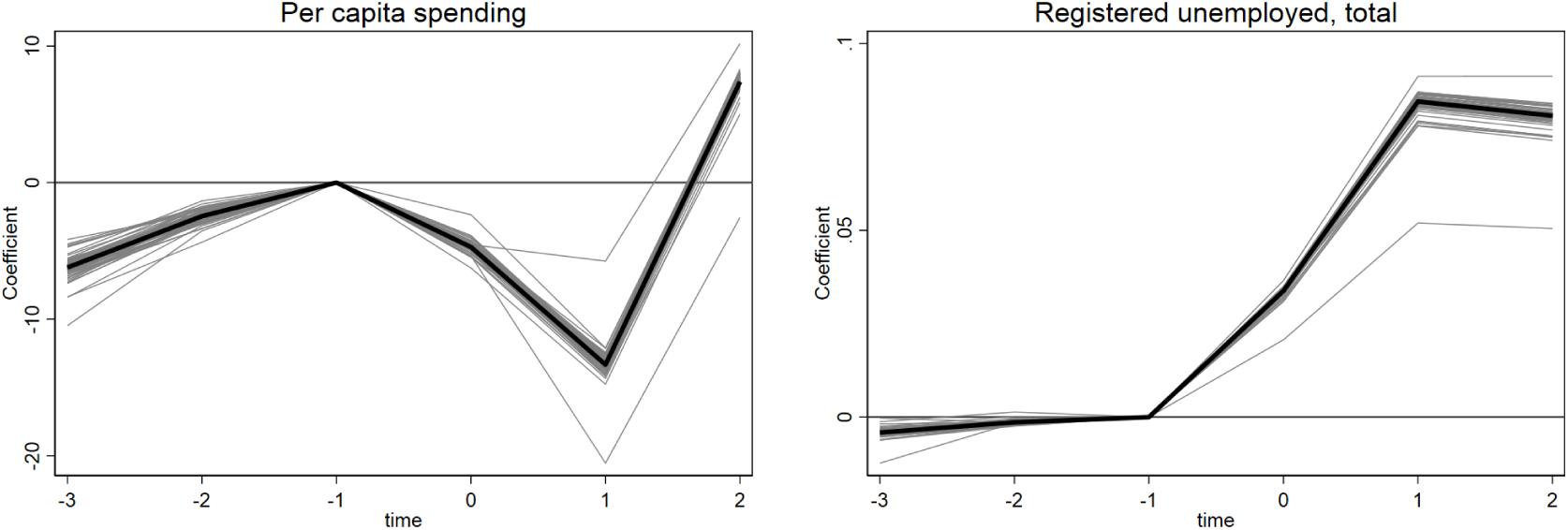
Robustness - Leave one out *Note:* This figure shows estimates of equation (1) with 95% confidence intervals leaving out one event. The specification that includes all events is marked as a black line. The estimates are population-weighted, and the standard errors are clustered at the municipality level.

**Table A6:** Summary statistics, by event window

|  | (1) | (2) | (3) | (4) | (5) | (6) |
| --- | --- | --- | --- | --- | --- | --- |
|  | 1+ mandate | No mandate | 1+ mandate | No mandate | 1+ mandate | No mandate |
| Incidence (per 100,000) | 8.834<br>(14.27) | 5.375<br>(9.042) | 8.284<br>(14.22) | 4.020<br>(8.843) | 8.569<br>(14.33) | 2.516<br>(8.083) |
| Hospitalizations (per 100,000) | 0.302<br>(1.285) | 0.185<br>(0.904) | 0.279<br>(1.355) | 0.128<br>(0.883) | 0.297<br>(1.375) | 0.0770<br>(0.929) |
| Tests (per 100) | 387.7<br>(255.8) | 280.6<br>(212.3) | 348.9<br>(237.2) | 252.8<br>(207.8) | 361.0<br>(251.0) | 223.7<br>(202.3) |
| Age | 40.16<br>(2.051) | 41.15<br>(2.494) | 40.46<br>(2.280) | 41.66<br>(2.515) | 40.28<br>(2.351) | 42.11<br>(2.386) |
| Foreign born | 0.178<br>(0.0563) | 0.154<br>(0.0620) | 0.177<br>(0.0655) | 0.136<br>(0.0538) | 0.183<br>(0.0697) | 0.120<br>(0.0323) |
| Higher ed | 0.392<br>(0.103) | 0.341<br>(0.102) | 0.381<br>(0.109) | 0.311<br>(0.0886) | 0.383<br>(0.109) | 0.284<br>(0.0611) |
| Cramped housing | 0.114<br>(0.0400) | 0.0993<br>(0.0420) | 0.113<br>(0.0454) | 0.0894<br>(0.0353) | 0.119<br>(0.0499) | 0.0792<br>(0.0176) |
| Low income | 0.116<br>(0.0242) | 0.108<br>(0.0241) | 0.114<br>(0.0268) | 0.103<br>(0.0209) | 0.116<br>(0.0279) | 0.0981<br>(0.0159) |
| Population (in 1000s) | 194.3<br>(235.5) | 125.0<br>(230.7) | 184.8<br>(266.5) | 72.89<br>(178.8) | 224.0<br>(294.6) | 19.10<br>(17.74) |
| N | 8736 | 2154600 | 9520 | 1730736 | 8820 | 1432550 |
| Window | 14 | 14 | 28 | 28 | 35 | 35 |
| Events | 312 | 312 | 170 | 170 | 126 | 126 |

Population-weighted averages. Each sample contains treated municipalities and clean controls in a symmetric window around the implementation of new NPIs. See section 2 for details on sample construction and variable definitions.

**Figure A9:**
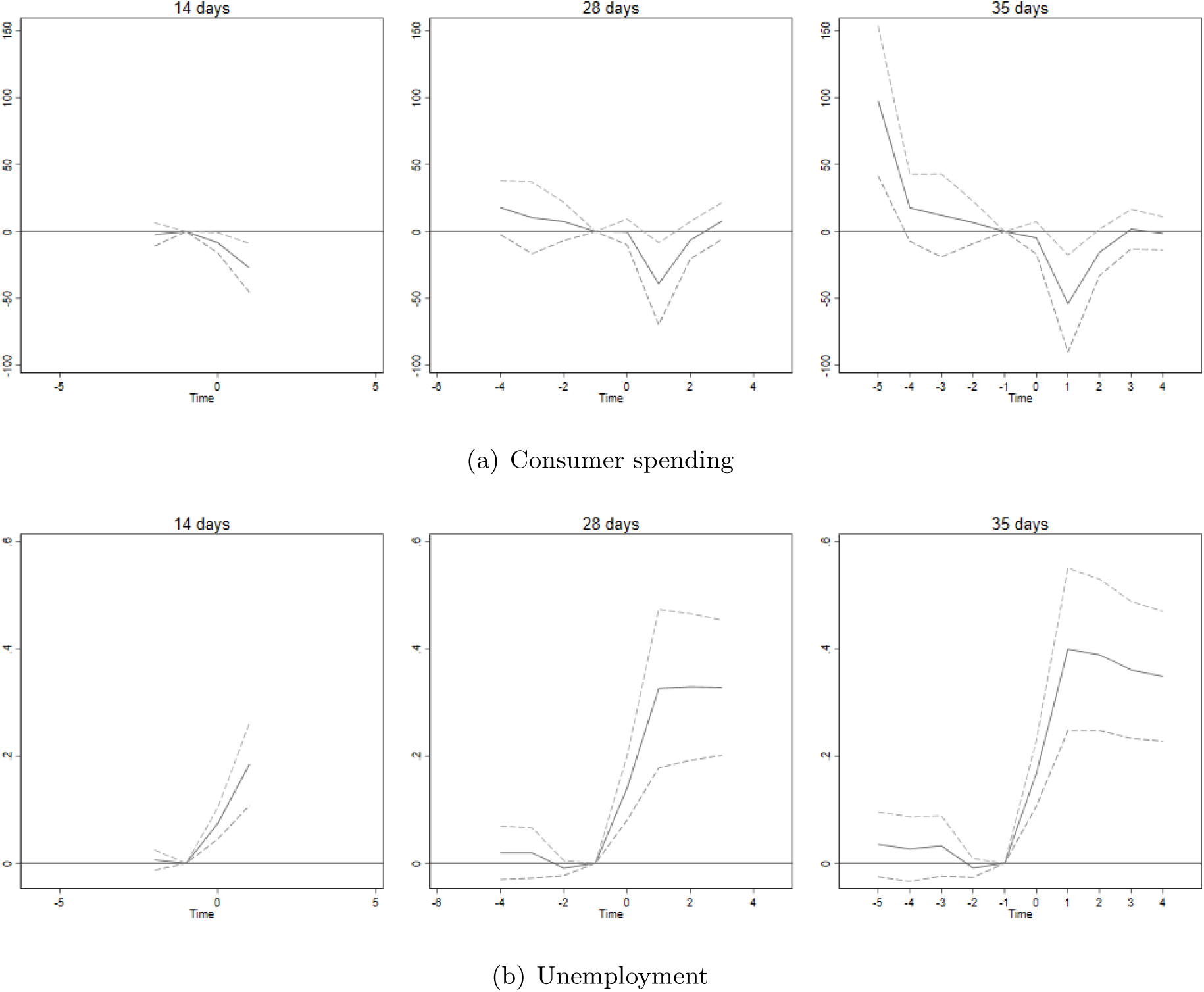
Event study - windows *Note:* This figure shows estimates of equation (1) for the different estimation windows with 95% confidence intervals. The estimates are population-weighted, and the standard errors are clustered at the municipality level.

**Table A7:** Policy simulations

| Outcome | Point estimate | Lower bound | Upper bound |
| --- | --- | --- | --- |
| Tests | -2938 | -6129 | 253 |
| Incidence | -369 | -538 | -200 |
| Hospitalizations | -10 | -18 | -2 |
| Registered unemployed, total | 10823 | 3561 | 18084 |
| Temp layoff, partial | 924 | -348 | 2196 |
| Temp layoff, full | 3265 | 1101 | 5428 |
| Ord unemployed, partial | 1178 | -420 | 2777 |
| Ord unemployed, full | 5456 | 2080 | 8832 |

*Note:* Table shows results from simple policy simulations showing estimated changes in the absolute number of tests, confirmed cases, hospitalization, and unemployment. Numbers obtained by multiplying the estimated $\hat{\rho}^{POST}$ from equation (2) by the total population in treated municipalities summed across events: $\Delta_{pol} = \sum_k treatpop_k \times \hat{\rho}_{pol}^{POST}$ . Lower and upper bound obtained by the lower and upper bounds of the 95% confidence intervals associated with $\hat{\rho}^{POST}$ .

**Figure A10:**
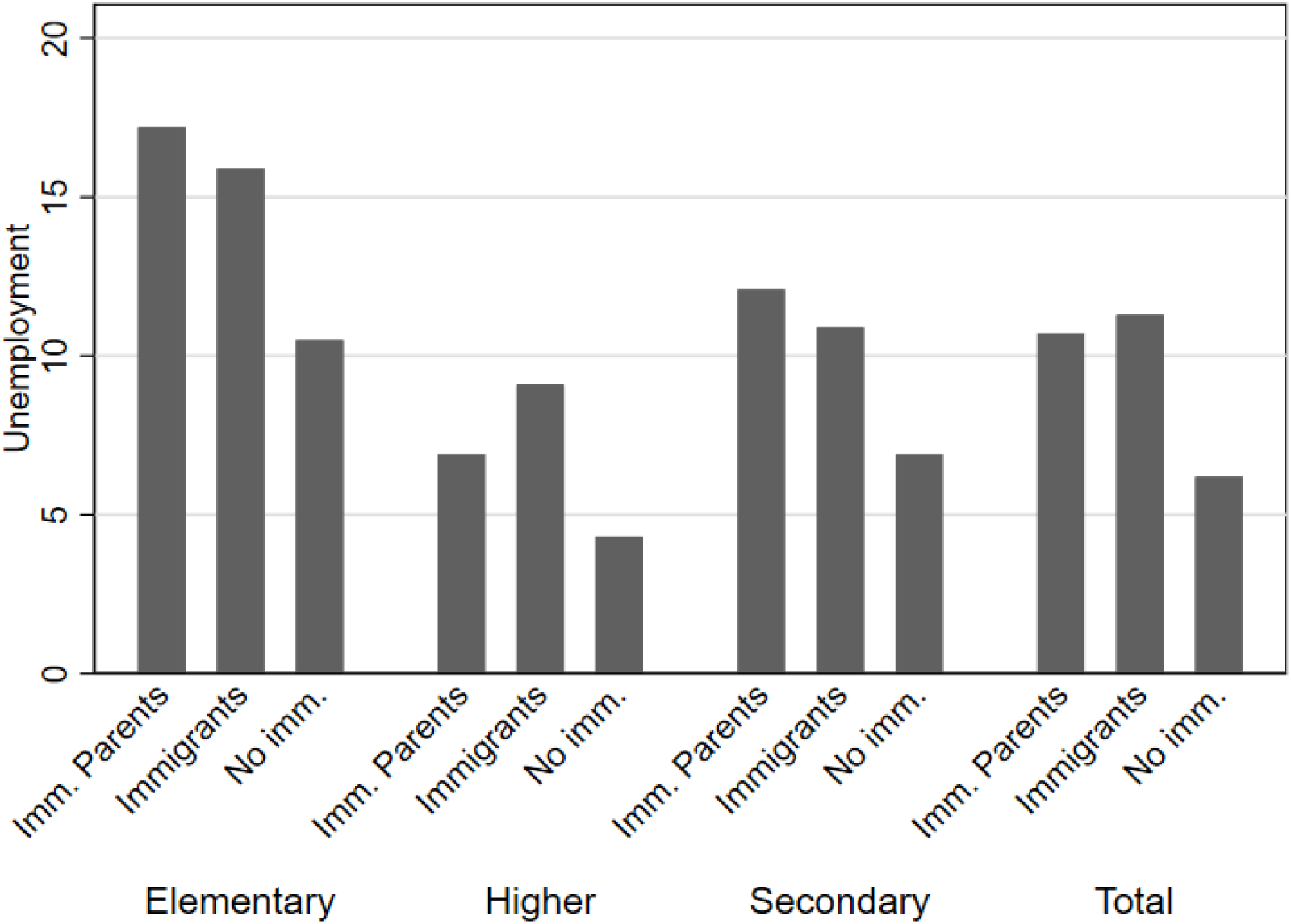
Unemployed during the pandemic by education and immigrant group *Note:* This figure shows the fraction of people who were employed in 2019, who were unemployed in the fourth quarter of 2020, by educational group and immigration background. Source: Statistics Norway, 2021.

## Footnotes

1 As we discuss in section 2, the Norwegian policy response stopped short of issuing blanket stay-at-home orders or curfews of the kind observed in several US states and European countries.

2 See https://www.vg.no/spesial/corona/tiltak/ for details. While local and regional policies have been an important part of the policy response from an early stage of the pandemic, there are currently no complete public databases documenting the timing and nature of these policies.

3 This restriction follows from our event study setup where we require an 6 week window around each event.

4 For hospitalizations, we use the date of the positive test.

5 The data were accessed through https://github.com/ActiveConclusion/COVID19 mobility.

6 The google mobility data contain other categories as well, but due to a non-trivial amount of missing data in some of these categories, they are left out of the analysis.

7 See figure 1 for trends in testing, incidence, and hospitalization for treatment and comparison municipalities.

8 Note that the Google mobility data is already a normalized measure of mobility relative to pre-pandemic baseline levels.

9 As the credit/debit card data is available only on a weekly basis (not daily), event time here is defined differently relative to the last full pre-implementation week. That is, event week 0 has between 1 and 7 post implementation days. This is different from in the models using daily data, where the NPIs are implemented on day 1 of week 0. Another issue is that there can be a few days delay in when payments are final and visible on the bank transfers.

10 While we do find a statistically significant increase in registered unemployment in neighboring municipalities the week of implementation, the effect is economically small (0.6% relative to pre-policy mean).

11 Common support graphs are presented in appendix figure A7.

12 See figure A9 in the appendix for event study graphs.

